# Established polygenic risk score for hypercholesterinemia demonstrates discriminatory value and risk stratification in an independent German Cohort

**DOI:** 10.64898/2026.09.01.26361969

**Authors:** Akhil Velluva, Linnaeus Bundalian, Erind Gjermeni, Julius L Katzmann, Ulrich Laufs, Ulrike Schatz, Stefan Bornstein, Rebecca Prielipp, Antje Garten, Johannes Schumacher, Rami Abou Jamra, Diana Le Duc

## Abstract

Polygenic risk scores (PRS) have emerged as promising tools for stratifying inherited disease risk, yet their translation into clinical practice is constrained by a critical and frequently unmet requirement: demonstration that scores derived in one cohort retain discriminatory value when applied independently in a different population. For suspected familial hypercholesterolemia (FH), a substantial proportion of patients that meet clinical criteria still test negative for a monogenic cause. These patients are presumed to carry polygenic LDL-C burden, yet PRS validation is still scarce. Here, we evaluated a published hypercholesterolemia PRS (PGS000936) spanning 5,386 genome-wide loci. We tested the score in 117 monogenic-negative hypercholesterolemia cases and 496 controls from the German general population genotyped on the Illumina Global Screening Array v3.0, with imputation to approximately 11 million variants. The score was applied without retraining, using externally derived β-coefficients. The PRS clearly distinguished cases from controls (p < 2×10⁻¹⁶), with a mean PRS of 0.97 in cases versus 0.52 in controls. Decile analysis revealed a seven-fold increase in odds of hypercholesterolemia in the top 10% of the distribution (OR 7.55; 95% CI 4.36–13.07; p < 0.0001). In 94 cases for which clinical data was available, higher PRS was associated with significantly higher LDL-C levels before treatment. Importantly, the PRS distribution in the German control cohort was shifted relative to that of the control cohort in the initial study, suggesting that population-matched calibration is required for accurate odds ratio estimation and proper interpretation of PRS values. These findings show that the published hypercholesterolemia PRS (PGS000936) can effectively stratify risk and identify individuals with a high genetic burden in a real-world German clinical setting, supporting its clinical utility when appropriately calibrated for the local population.

## Introduction

The prospect of using an individual’s genome to anticipate disease risk has long been a central ambition of human genetics. Polygenic risk scores (PRS) represent the most mature realization of this ambition for common complex diseases. By aggregating the small, additive effects of thousands of common single-nucleotide polymorphisms (SNPs) identified through genome-wide association studies (GWAS), PRS distil a complex polygenic architecture into a single continuous measure of inherited susceptibility [1,2]. As GWAS sample sizes have expanded into hundreds of thousands and even millions of participants, the discriminatory power of PRS has increased substantially, and scores are now available for hundreds of clinically relevant traits [3]. This has fueled growing interest in translating PRS from research tools into clinical instruments, with proposed applications spanning cardiovascular disease, metabolic disorders, psychiatric conditions, and cancer screening [2,4,5].

Despite this momentum, the path from PRS derivation to clinical implementation is not straightforward. A fundamental and often underappreciated challenge is transportability: whether a score developed in one population retains its discriminatory performance and calibration when transferred to an independent cohort. Most large-scale PRS have been derived in European-ancestry biobanks, most prominently the UK Biobank (n > 450,000), and their performance characteristics are inherently shaped by the linkage disequilibrium (LD) structure, allele-frequency spectrum, ascertainment design, phenotype definition, and environmental background of that source population [6,7]. When transferred to an independent cohort that differs in any of these dimensions, predictive performance can degrade substantially, a phenomenon well-documented across diverse ethnic groups, disease areas, and clinical contexts [6,8]. Crucially, poor calibration may not be apparent from the score itself and can only be detected by explicit comparison with a local, population-matched control distribution. Without this step, direct import of source-study risk thresholds will produce systematically misleading risk estimates.

The need for robust PRS validation is highly relevant in the context of familial hypercholesterolemia (FH). FH has traditionally been considered a monogenic disorder caused by pathogenic variants in genes such as *LDLR*, *APOB*, or *PCSK9*. However, a substantial proportion of individuals who fulfill clinical diagnostic criteria for FH do not harbor an identifiable disease-causing variant despite comprehensive genetic testing [9]. Multiple lines of evidence now support the concept of polygenic hypercholesterolemia as a biologically and clinically distinct entity: individuals without a monogenic FH mutation often carry a significantly elevated burden of common LDL-C-raising alleles compared with population controls, consistent with the additive accumulation of many small-effect variants rather than a single large-effect mutation [10–12]. The distinction between monogenic and polygenic FH has practical relevance: monogenic carriers face a higher lifetime cardiovascular risk and a greater likelihood of an affected first-degree relative, whereas polygenic cases may have a somewhat more moderate risk trajectory, with implications for treatment intensity and cascade testing strategies [11,13].

Consequently, PRS offers a valuable framework for distinguishing monogenic from polygenic forms of hypercholesterolemia, refining genetic diagnoses, improving risk stratification, and informing clinical management and cascade screening strategies. Nonetheless, both groups benefit from early lipid-lowering intervention.

Because these applications depend on accurate classification of individuals relative to their background population, proper calibration and validation of hypercholesterolemia PRS within local clinical cohorts are essential before routine implementation.

In this study, we address this gap. Our primary aim was to determine whether an externally derived, publicly available hypercholesterolemia PRS, applied without any retraining or coefficient modification, can distinguish clinically ascertained monogenic- negative cases from German population controls. Secondary aims were to quantify risk enrichment across PRS strata, assess calibration relative to the source cohort, test whether PRS associates with the severity of LDL-C elevation in cases with available clinical data before treatment. We frame this work explicitly as an external validation and implementation-focused analysis rather than a PRS construction exercise, because demonstration of real-world performance in an independent clinical setting is now one of the most critical and insufficiently addressed needs in the field [2,8].

## Materials and Methods

### The UK Biobank cohort study

The score definitions were fixed prior to analysis and were not modified, retrained, or reweighed at any stage. The tested score was PGS000936, a sparse genome-wide model comprising 5,386 loci derived from UK Biobank-based GWAS analyses and reported by Tanigawa *et al*. [3]. We selected this PRS because it was constructed from a large, well-powered discovery dataset using established penalized regression methods that prioritize a sparse but well-calibrated set of variants, and because it is publicly archived in the PGS Catalog with fully documented β-coefficients and allele specifications.

#### Genotyping and Quality Control

All 613 samples (117 cases, 496 controls) were genotyped using the Illumina Global Screening Array v3.0, a high-density BeadChip platform optimized for genome-wide association and polygenic risk score applications. This array provides comprehensive coverage of common variants across the genome and is widely used in European population-based studies. Genotype calling was performed using Illumina GenomeStudio software with export in PLINK-compatible format. All genomic positions were referenced to GRCh37 (hg19). Quality control was implemented following published best-practice recommendations for Illumina genotyping arrays [14]. At the sample level, exclusions were applied for low call rate, excess heterozygosity (indicative of DNA quality problems or sample contamination), and discordance between genotypic and reported sex. At the variant level, SNPs with a call rate below 95% or a minor allele frequency (MAF) below 1% were excluded. These thresholds are standard in GWAS and PRS quality control pipelines and minimize the contribution of low-confidence genotype calls to downstream score construction.

#### Strand Alignment and Variant Position Validation

Accurate strand orientation is essential for PRS construction because β-coefficients from source studies are anchored to specific effect alleles on a defined strand. Strand- flip errors, in which the complement allele is inadvertently coded as the effect allele, can reverse the direction of a SNP’s contribution to the PRS and introduce systematic bias. To address this, strand-flip correction was applied to all variants using an in-house pipeline referencing the 1000 Genomes Project phase 3 haplotype panel [15]. Variant positions were independently validated against the GRCh37 reference assembly using dbSNP annotations and internal consistency checks. Variants with ambiguous strand orientation (A/T and C/G SNPs at intermediate MAF) were excluded to prevent misalignment that cannot be resolved by strand flip alone [16].

#### Genotype imputation and quality control

Because array-based genotyping captures only a fraction of common variants genome-wide, imputation was performed to expand genomic coverage and maximize overlap between the target dataset and the external PRS variant lists. Imputation was carried out using Beagle version 5.4, a widely validated framework for haplotype phasing and genotype imputation from array data [17]. Each chromosome was processed independently on a high-performance computing node using 24 parallel cores and 12 iterations per chromosome, resulting in runtimes of approximately 2–3 hours per chromosome. Reference haplotypes were drawn from the 1000 Genomes Project phase 3 panel and genetic map positions from the HapMap phase II map. Following imputation, the dataset contained approximately 11 million variants genome-wide, substantially extending coverage beyond the ∼700,000 positions directly typed on the array.

The choice of Beagle 5.4 is supported by benchmark comparisons demonstrating high imputation accuracy for European-ancestry samples, particularly for common and low-frequency variants (r² > 0.8 for MAF > 5%) when reference panels of comparable size are used [17]. The 1000 Genomes Project European subset (503 individuals across 5 populations) was used as the primary reference rather than larger panels such as the Haplotype Reference Consortium (n = 64,976) because of its public accessibility and the near-perfect allele-frequency concordance it achieves with European-ancestry study cohorts at MAF > 1% [15,18].

Because poorly imputed variants can introduce noise and spurious associations into PRS calculations, a stringent post-imputation quality filter was applied. Variants were retained only if they achieved an R² metric of ≥ 0.5, where R² reflects the squared correlation between imputed and true genotypes as estimated by Beagle’s internal algorithm. This threshold is widely used in imputation-based analyses and balances retention of informative variants against exclusion of unreliably imputed loci [19]. To validate the quality of the retained imputed variants empirically, allele frequencies at qualifying loci were compared with those from the European subset of the 1000 Genomes Project, stratified by genotype class (homozygous reference 0|0, homozygous alternative 1|1, and both heterozygous classes 1|0 and 0|1). High concordance between imputed frequencies and reference frequencies across all genotype classes (Figure 1) was taken as evidence that imputation had not introduced systematic misclassification that could bias downstream PRS construction [15,17].

**Fig. 1.**
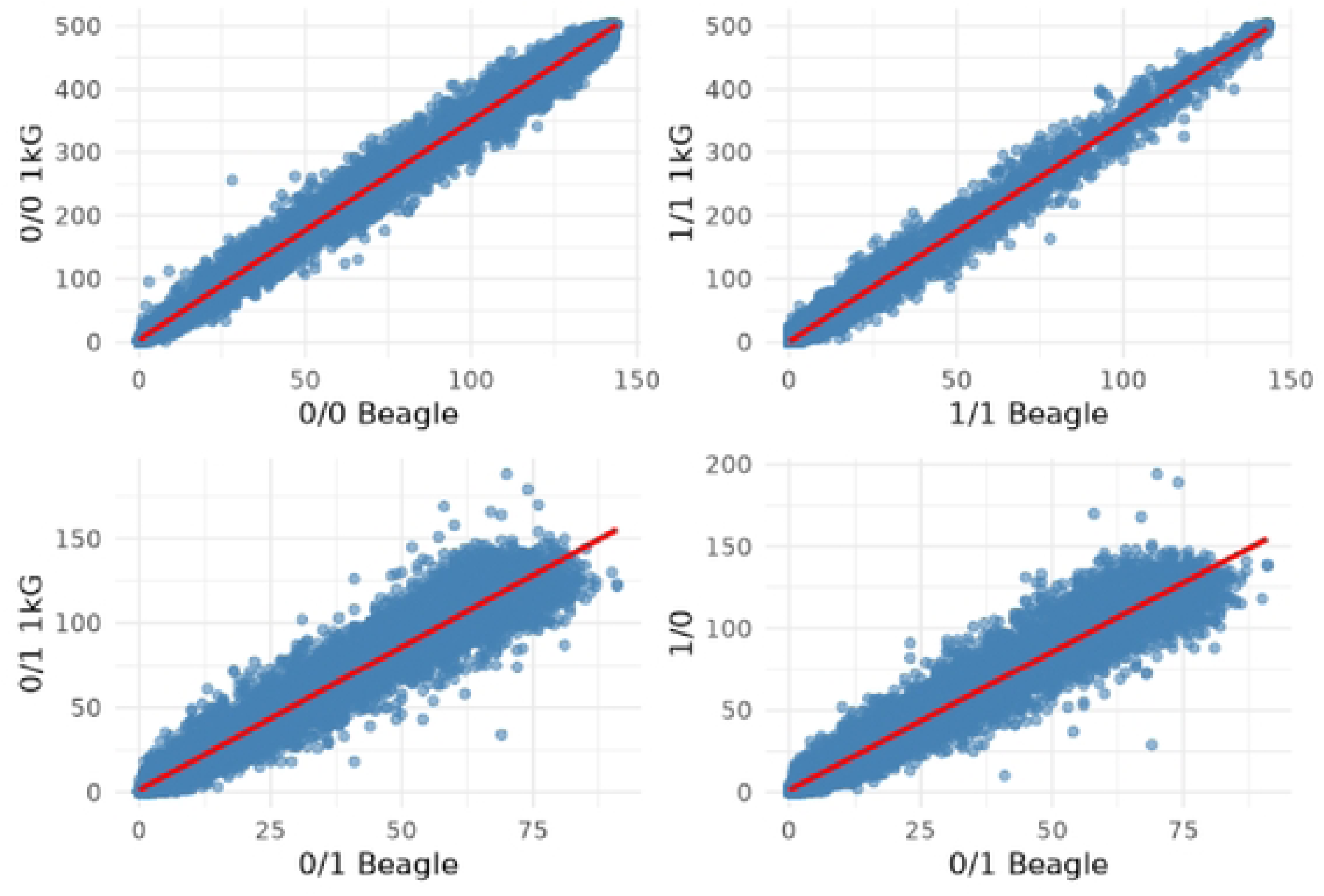
Allele Frequency Correlation of European Population and Imputed data. The high correlations observed reflect the close genetic background shared between the two populations – imputed and European reference population, particularly for common variants.

#### Polygenic Risk Score Calculation

PRS were calculated under the standard additive model, which assumes that the contribution of each locus to the phenotype is proportional to the number of effect alleles carried, scaled by the externally derived effect size βⱼ [2]. For individual i at locus j, the score contribution was βⱼ × Gᵢⱼ, where Gᵢⱼ ∈ {0, 1, 2} denotes the dosage of effect alleles. Homozygous effect-allele carriers (1|1) contributed 2β, heterozygous carriers (0|1 or 1|0) contributed β, and non-carriers contributed 0. The individual-level PRS was the sum of these contributions across all qualifying loci. For the primary PGS000936 score, 4,240 of the 5,386 score loci were represented by imputed variants and 1,146 by directly genotyped variants. For the 223-SNP secondary score, all loci present in the imputed dataset after quality filtering were included. This approach follows the β- transfer framework: the reuse of externally derived effect sizes in an independent target cohort without recalibration, which is valid when the discovery and target cohorts share comparable ancestry and phenotype definitions [2,6].

#### Statistical Analysis

Case-control differences in PRS distributions were assessed using the Welch two-sample t-test, which does not assume equal variances and is appropriate for comparing distributions of differing sample sizes. To characterize the gradient of risk enrichment across the score range, individuals were grouped into PRS deciles and odds ratios (ORs) with 95% confidence intervals were computed for each stratum relative to the lowest decile using logistic regression. This decile-stratified analysis is a standard approach in PRS validation literature and enables direct assessment of whether the score provides clinical utility beyond a simple mean difference [20,21]. In the subset of 94 cases with available clinical data, the association between PRS strata and the highest documented untreated LDL-C concentration was tested using linear regression adjusted for age and sex. PRS was modeled both as quintiles and deciles to assess consistency of the association across binning strategies. All analyses were conducted in R (version 4.3). Statistical significance was defined as p < 0.05 (two-tailed).

#### Ethics statement

The study was approved by the Ethics Committee at Leipzig University (approval no. 402-16ek), and all participants provided written informed consent for genetic analyses and the use of their pseudonymized data for research into the genetic basis of disease, including secondary research analyses and reporting of results at an aggregate level. In participants with a clinical suspicion of familial hypercholesterolemia in whom no monogenic cause was identified, polygenic risk score (PRS) analysis was subsequently performed as an exploratory secondary analysis. The data was accessed for the analysis in 2024, and the authors did not have access to any information that could directly identify individual participants. PRS results were evaluated for research purposes only and are reported exclusively at the aggregate cohort level; no individual PRS results were used for clinical decision-making or returned to participants.

## Results

### Genotype imputation closely mirrors the allele frequency distribution of European reference population

Post-imputation quality control retained variants with R² ≥ 0.5, providing a set of well- supported loci for PRS construction. To empirically verify the accuracy of imputation in our cohort, allele frequencies at retained imputed loci were compared with those from the European subset of the 1000 Genomes Project, stratified by genotype class. Concordance was consistently high: Pearson correlation coefficients were 0.99 for homozygous reference (0|0) and homozygous alternative (1|1) genotypes, and 0.97 for both heterozygous classes (1|0 and 0|1) (Table 1). These correlations indicate near- perfect agreement between imputed genotype frequencies and the European reference population for common and well-imputed variants and confirm that systematic imputation artifacts are unlikely to have biased downstream score calculations. The high concordance across all four genotype classes, including the heterozygous calls that are inherently more uncertain than homozygotes, is consistent with benchmark performance reported for Beagle 5.4 in European-ancestry cohorts and provides confidence in the reliability of the imputed data for PRS construction [17].

**Table 1.** Allele Frequency Correlation of European Population and Imputed data. The is a high correlation between our study cohort and the European reference population.

| Genotype | Correlation coefficient |
| --- | --- |
| 0 \| 0 | 0.99 |
| 1 \| 1 | 0.99 |
| 1 \| 0 | 0.97 |
| 0 \| 1 | 0.97 |

### The tested polygenic risk score discriminates disease susceptibility and stratifies risk by case-control contrasts

Application of the PGS000936 score to our German cohort produced a marked and statistically unambiguous separation between hypercholesterolemia cases and controls (Figure 2). The mean PRS was 0.97 (SD 0.42) in cases compared with 0.52 (SD 0.40) in controls, a difference of approximately 1.1 standard deviations (Welch t-test, p < 2×10⁻¹⁶). This result demonstrates that an externally derived score, built in a UK Biobank population using UK-specific LD structure and allele frequencies, retains strong discriminatory value when transferred without modification to a clinically ascertained German cohort. The magnitude of separation is notable given that the score was not derived or optimized in any German-specific dataset and was applied to a relatively small cohort of 613 individuals, circumstances that typically attenuate PRS performance compared with large biobank validation studies. The observed discrimination is therefore likely to represent a conservative estimate of the score’s performance with larger sample sizes.

**Fig. 2.**
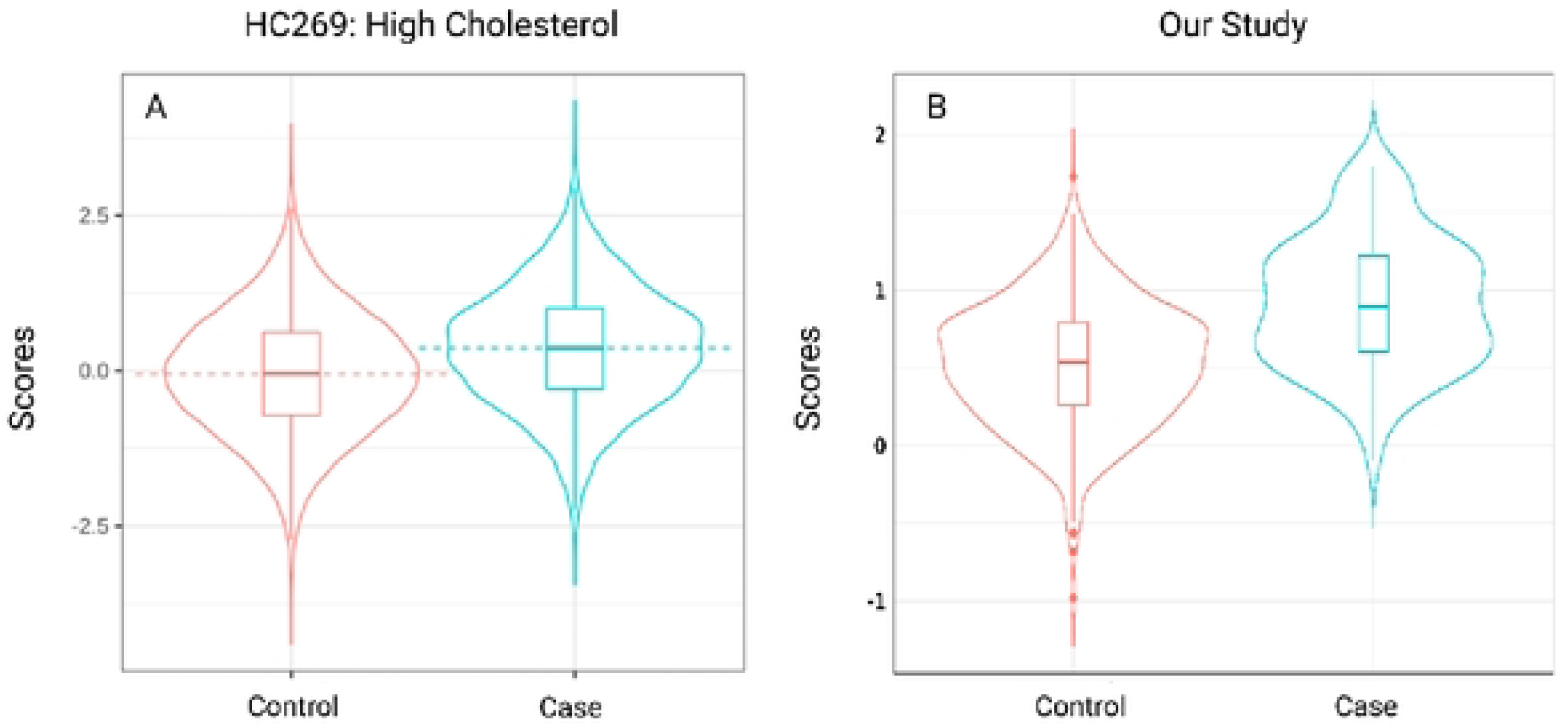
Comparison of polygenic risk score (PRS) distribution of 2 related studies A) PRS score distribution for high cholesterol cases in the original study B) PRS score distribution calculated for a hypercholesteremia cohort in the current study. PRSs were calculated using published β-coefficients from PGS catalog PGS000936 (5,386 SNPs; 4,240 imputed, 1,146 genotyped). A. The mean PRS was significantly higher in cases than in controls (Welch t-test, p < 2e-16), demonstrating the predictive value of well-calibrated PRSs. B. Compared to the original study, the control distribution in our cohort had a mean >0, indicating that direct comparison to the original cases would overestimate risk and highlight the need for population- matched calibration.

A critically important observation emerged from comparison of the score distributions with those of the source study. In the original UK Biobank-derived analysis, the control PRS distribution was centered near zero, as expected for a score calibrated such that the population mean represents the reference level. In our German cohort, however, the control distribution was shifted rightward, with a mean PRS substantially above zero (Figure 2). This shift demonstrates that the score’s absolute values are not universal and depend on the ancestry composition, ascertainment, and environmental background of the target cohort. Directly applying the source study’s risk thresholds or percentile cutoffs to our cohort would have produced systematically inflated risk estimates, a finding with direct implications for any clinical deployment of PRS. Odds ratios and risk strata must therefore be recalibrated using controls from the target population. Importantly, this calibration does not require a very large sample: while deriving reliable β-coefficients requires massive discovery cohorts, meaningful calibration of an existing score requires only a modest-sized, population-matched reference distribution[2,8].

### Risk enrichment is strikingly concentrated in the upper decile of the PRS distribution

Stratification of cases and controls into PRS deciles revealed a pronounced and clinically compelling pattern of risk enrichment (Figure 3). The relationship between PRS and hypercholesterolemia odds was not uniform across the distribution: the most striking effect was confined to the uppermost stratum. Individuals in the top 10% of the PRS distribution had an odds ratio of 7.55 (95% CI: 4.36–13.07; p < 0.0001) compared with the lowest decile. This translates into a seven-fold increase in the odds of presenting clinically significant hypercholesterolemia even in the absence of a monogenic FH mutation, a magnitude of effect that is clinically substantial and statistically robust. In contrast, the second-highest stratum showed a considerably attenuated effect (OR 1.80; 95% CI: 1.03–3.14; p = 0.038), and intermediate strata exhibited non-significant odds ratios (OR 1.06; 95% CI: 0.58–1.95; p = 0.86).

**Fig. 3.**
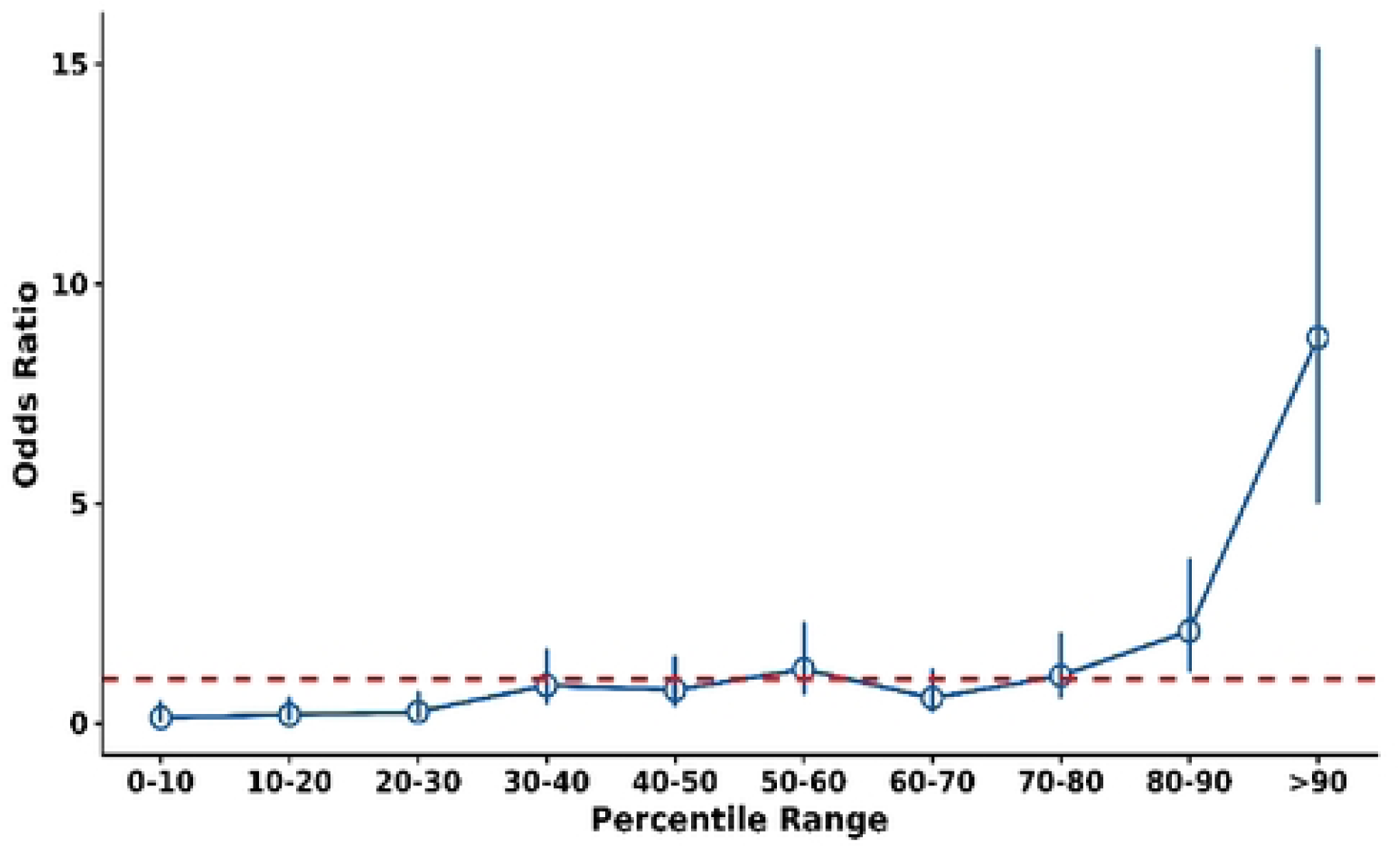
PRS decile-stratified odds ratios for hypercholesterolemia. Odds ratios were observed to increase at upper end of the PRS distribution. With OR of 7.55 (95% CI: 4.36-13.07; p < 0.0001) in the highest decile (>90%).

**Fig. 4.**
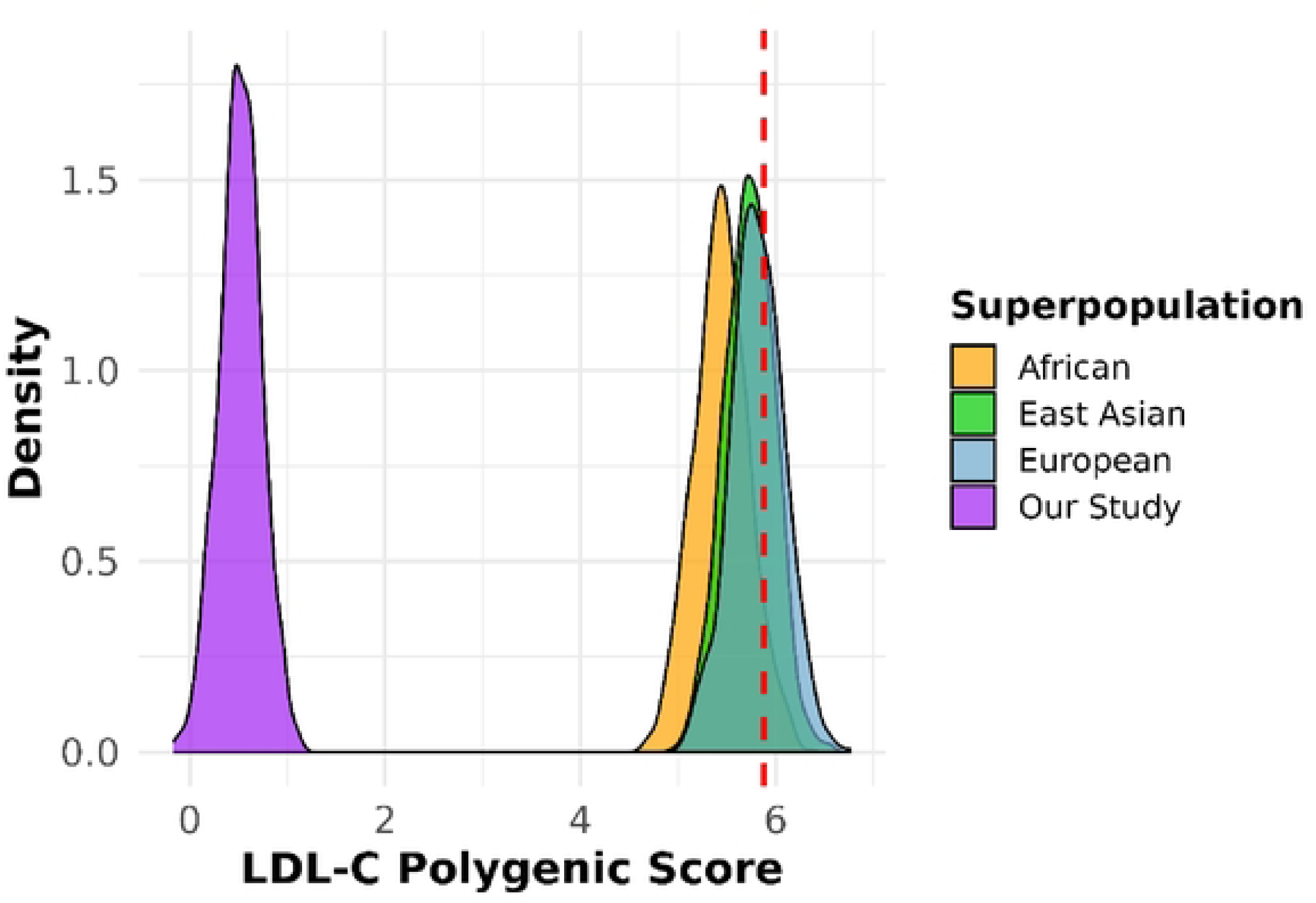
Calculated PRS distrubution using β-coefficients for 223-SNPs.

This non-linear enrichment pattern, strong signal at the tail, weak signal in the middle, is consistent with patterns reported in cardiovascular PRS literature more broadly. Studies of LDL-C polygenic scores in general population cohorts have similarly shown that the strongest clinically actionable signal resides in the upper 5–10% of the polygenic score distribution, where lifetime LDL-C burden is disproportionately elevated and cardiovascular event rates are substantially higher than in individuals at average genetic risk [20,21]. The finding that this same architecture is preserved in our smaller, clinically ascertained cohort, where all cases have already crossed a clinical threshold for LDL-C elevation, reinforces the robustness of the signal. It also has direct implications for clinical use: the primary utility of PRS in the FH referral setting is not to assign numerical probabilities to all patients, but to identify the subset at the uppermost extreme of polygenic burden who may warrant the most intensive clinical attention.

### Higher PRS strata are associated with greater untreated LDL-C concentrations in clinically characterized cases

Extended clinical data was available for 94 of the 117 cases (Table 2). This subset was predominantly female (58.5%), with a mean age of 52.6 years (SD 12.3), reflecting the age and sex distribution of individuals referred to lipid clinics for hypercholesterolemia evaluation. The cardiovascular risk profile of this cohort was substantial: coronary artery disease was present in 22.3% of patients, myocardial infarction in 17.0%, and hypertension in 47.9%. Two-thirds of patients (67.0%) had a first-degree relative with elevated LDL-C, consistent with a familial clustering pattern. The mean highest documented untreated LDL-C concentration was 6.5 mmol/L (SD 1.2), confirming that these individuals had markedly elevated LDL-C at presentation, with levels well above the guideline-based threshold for FH diagnosis (≥5.0 mmol/L).

**Table 2.** Baseline characteristics of n=94 with available clinical data. The discriminating power of PRS observed through case-control contrasts and stratified risk gradients at the population level provides robust empirical evidence supporting the feasibility of cross-cohort adaptation of β effect sizes or “β transfer” for reliably informing risk models in during cross-cohort adaptation.

|  |  |
| --- | --- |
| N | 94 |
| Female (%) | 58.5 |
| Age (years [SD]) | 52.6 (12.3) |
| Highest untreated LDL-C (mean in mmol/L [SD]) | 6.5 (1.2) |
| Lowest LDL-C under treatment (mean in mmol/L [SD]) | 2.0 (1.1) |
| Coronary artery disease (%) | 22.3 |
| Myocardial infarction (%) | 17.0 |
| Stroke or TIA (%) | 2.1 |
| Peripheral artery disease (%) | 5.3 |
| Hypertension (%) | 47.9 |
| Diabetes mellitus (%) | 5.3 |
| Statin treatment at inclusion (%) | 35.1 |
| First degree relative with elevated LDL-C (%) | 67.0 |
| First degree relative with premature cardiovascular event (%) | 17.0 |
| Xanthoma (%) | 1.1 |
| Corneal arcus < 45 years (%) | 0 |
| Xanthelasma (%) | 5.3 |

Within this clinically characterized subset, PRS strata were significantly associated with the highest documented untreated LDL-C in linear regression models adjusted for age and sex (p = 0.033 for quintiles; p = 0.041 for deciles). This finding is conceptually important because it extends the score’s utility beyond a binary case-control comparison. The fact that PRS tracks quantitative LDL-C severity within a cohort of cases who are already phenotypically extreme indicates that the score captures genuine variation in inherited polygenic LDL-raising burden, rather than simply reflecting a threshold effect at the case-control boundary. This dose-response relationship between polygenic score and LDL-C phenotype is consistent with a large body of Mendelian randomization and polygenic burden literature demonstrating that LDL-C genetic scores track continuous lipid concentrations across the full population distribution [20,22], and its preservation within a mutation-negative clinical cohort further validates the biological coherence of the PRS signal.

### Variant-level analysis reveals the relative contribution of genetic burden

Inspection of variant-level score contributions across PRS deciles revealed biologically coherent patterns that support the validity of the observed PRS signal. Individuals in the highest PRS deciles carried a disproportionate burden of risk alleles at loci with well-characterized roles in LDL metabolism, most prominently *APOB*, *LDLR*, and *PCSK9*. These three genes encode canonical components of the LDL receptor pathway: LDLR mediates hepatic LDL particle uptake; APOB is the principal apolipoprotein of LDL particles and the ligand for LDLR; and PCSK9 regulates LDLR recycling through proteasomal degradation [23]. Pathogenic variants in all three genes cause monogenic FH; common variants in the same genes and their regulatory regions are among the strongest contributors to population-level LDL-C variation identified in GWAS [24]. The enrichment of common risk alleles at these canonical loci in higher-scoring individuals is precisely what would be expected if the PRS is capturing genuine inherited polygenic LDL-C burden rather than noise or technical artifact, and it provides biological plausibility for the statistical signal observed in case-control analysis.

## Discussion

This study provides an empirically grounded demonstration that an externally derived, publicly available hypercholesterolemia PRS retains strong discriminatory performance when applied to an independent German cohort of clinically ascertained, monogenic- negative patients, without any score retraining or coefficient modification. The primary findings are: (i) the PGS000936 score produced a highly significant separation between cases and controls; (ii) risk enrichment was strikingly concentrated at the upper end of the distribution, with a seven-fold increase in hypercholesterolemia odds in the top decile; (iii) PRS strata tracked untreated LDL-C concentrations within the clinical case cohort; and (iv) local calibration against population-matched controls is essential because the absolute score distribution is not transferable on a universal scale. Together, these results establish that published PRS can add genuine clinical value in a real-world German FH referral setting and provide a model for how such validation should be conducted.

The clinical context of this cohort is fundamental to the significance of these findings. Mutation-negative suspected FH represents a diagnostically challenging and clinically pressing issue in lipid medicine. Patients in this group present with severe LDL-C elevation, a family history of hypercholesterolemia, and elevated cardiovascular risk, yet standard genetic testing provides no actionable result. In this diagnostic vacuum, clinicians are left without a molecular explanation for the phenotype and without a basis for communicating genetic risk to the patient or to at-risk relatives. The concept of polygenic hypercholesterolemia, in which the phenotype reflects the cumulative effects of numerous common variants with small individual effect sizes rather than a single causative mutation, provides a biologically plausible explanation for these cases. However, its clinical utility depends on the availability of validated and readily deployable tools to quantify the underlying polygenic burden [10–12]. Our findings demonstrate that a freely available, externally validated PRS can provide such a tool, enabling quantitative assessment of polygenic burden and effective risk stratification in this diagnostically challenging population.

The decile analysis is the most clinically actionable finding in this study. A seven-fold increase in hypercholesterolemia odds in the top PRS decile is a large effect by the standards of common disease genetics, where even well-validated PRS typically yield odds ratios of 2–4 per standard deviation of score [2,20]. The concentration of this effect at the upper tail, with intermediate strata showing weak or non-significant enrichment, is consistent with broader cardiovascular PRS literature. Studies of LDL-C and coronary artery disease polygenic scores in population cohorts have consistently found that the clearest clinical signal resides in the top 5–10% of the distribution, where lifetime cumulative LDL-C exposure is disproportionately elevated and rates of premature atherosclerotic events are substantially higher [20,21]. In our study, this pattern is recapitulated in a clinically ascertained cohort where all cases already carry a high-risk phenotype. The practical implication is clear: the most productive use of PRS in the FH clinic is not to calculate a risk number for every patient, but to identify the subset in the top decile who carry the greatest inherited burden and who may warrant intensified treatment targets, more aggressive LDL-C monitoring, and targeted cascade screening of first-degree relatives.

The association between PRS strata and untreated LDL-C concentrations in the clinical subset reinforces this interpretation by anchoring the score to a quantitative, physiologically meaningful phenotype. Prior work on polygenic LDL-C scores in FH cohorts has shown that mutation-negative patients with elevated polygenic scores tend to have higher LDL-C than those with average scores, consistent with a dose- response relationship between polygenic burden and lipid concentration [10,11]. Our finding that this relationship persists after adjustment for age and sex in a German clinical cohort extends this evidence to a new population and to a new, larger score architecture. It also adds a dimension of clinical validation that goes beyond case-control discrimination: a score that tracks LDL-C severity within an affected group is demonstrating biological coherence, not merely statistical separation at the case-control boundary.

The calibration findings are equally important from an implementation perspective. The rightward shift of our German control distribution relative to the UK Biobank source study is not a sign of score failure; the score discriminated excellently despite this shift. Rather, it is a sign that absolute PRS values are population- and cohort-specific and cannot be used interchangeably across settings without recalibration. This point is increasingly emphasized in PRS portability and implementation literature, where calibration failure, rather than discrimination failure, has been identified as the primary obstacle to clinical translation[6,8]. Calibration failure occurs when the score’s absolute values are offset in the target cohort relative to the source, such that source-study percentile thresholds do not correspond to the same risk levels in the target population. In our cohort, using the source study’s percentile thresholds to define high-risk individuals would have classified an inflated proportion of controls as high-risk, leading to overdiagnosis and potentially unwarranted clinical intervention. The solution, recalibrating the score distribution against local controls, is straightforward and does not require large sample sizes, but it does require that institutions collect a local reference distribution before deploying externally derived PRS.

The variant-level enrichment of risk alleles at *APOB*, *LDLR*, and *PCSK9* in the highest PRS deciles provides biological credibility that the score is measuring what it claims to measure. These three genes define the LDL receptor pathway and are the canonical substrates of both monogenic FH and common LDL-C variation [23,24]. The fact that polygenic accumulation of common risk alleles at these same loci—in individuals who lack a monogenic mutation, tracks with hypercholesterolemia case status and LDL-C severity is consistent with the broader concept of a genetic continuum for LDL-C risk, in which monogenic and polygenic forms of FH represent the tail and the bulk, respectively, of a single underlying genetic distribution [10,25]. Our findings thus not only validate the PRS in a statistical sense but situate it within a biologically coherent framework that should be communicable to clinicians and patients alike.

Several limitations should be acknowledged. The sample size is modest by the standards of discovery-phase PRS studies: 117 cases and 496 controls provide adequate power to detect the large effects observed in the top decile but less power for precise estimation in intermediate strata. Although imputation quality was high and allele-frequency concordance with the European reference panel was excellent, array-based imputation cannot fully recover all variants in the score’s original derivation dataset, and missing variants may attenuate performance below its theoretical maximum. The study was also designed as a fixed external validation rather than an optimization exercise; a score retrained on German-specific data or on a larger European reference panel might achieve higher discrimination. Finally, while our controls were not suspected of having FH, they belonged to a cohort of psychiatric disorders.

In conclusion, this study makes three main contributions to the PRS implementation literature. First, it demonstrates that a publicly available, externally derived hypercholesterolemia PRS retains clinically meaningful discriminatory performance in an independent German cohort without any score modification, establishing that β- transfer is feasible across cohorts with comparable European ancestry and well-harmonized phenotype definitions. Second, it shows that the most informative signal is concentrated at the upper tail of the PRS distribution, with a seven-fold increase in hypercholesterolemia odds in the top decile, directly supporting the use of PRS to identify a high-burden subgroup within the diagnostically challenging mutation-negative FH population. Third, it provides a clear practical illustration of why local calibration with population-matched controls is mandatory for clinically meaningful interpretation, and why different externally derived scores for the same phenotype cannot be assumed to transfer equivalently. Together, these findings support the responsible integration of externally derived PRS into the clinical genetics workup of mutation-negative suspected FH, with the expectation that such scores will be most impactful when used to identify individuals at the extremes of polygenic burden rather than to assign absolute risks across the full distribution.

## Data Availability

The datasets used in this study can be found in https://www.ukbiobank.ac.uk/ and https://www.pgscatalog.org/ under the study name PGS000936.

https://www.pgscatalog.org/score/PGS000936/

## Acknowledgement

We thank the German Research Foundation and University of Leipzig for the opportunity for entrusting the grants to support the projects in Le Duc Lab.

